# Rapid homogeneous assay for detecting antibodies against SARS-CoV-2

**DOI:** 10.1101/2020.11.01.20224113

**Authors:** Juuso Rusanen, Lauri Kareinen, Lev Levanov, Sointu Mero, Sari H. Pakkanen, Anu Kantele, Fatima Amanat, Florian Krammer, Klaus Hedman, Olli Vapalahti, Jussi Hepojoki

**Affiliations:** Department of Virology, Medicum, Faculty of Medicine, University of Helsinki, Helsinki, Finland; Human Microbiome Research Program, Faculty of Medicine, University of Helsinki, Helsinki, Finland; Meilahti Vaccination Reasearch Center, MeVac, Inflammation Centre, Helsinki University Hospital and University of Helsinki, Helsinki, Finland; Department of Microbiology, Icahn School of Medicine at Mount Sinai, NY, USA; Graduate School of Biomedical Sciences, Icahn School of Medicine at Mount Sinai, NY, USA; HUS Diagnostic Center, HUSLAB, Helsinki University Hospital and University of Helsinki, Helsinki, Finland; Department of Veterinary Biosciences, Faculty of Veterinary Medicine, University of Helsinki, Helsinki, Finland; Institute of Veterinary Pathology, Vetsuisse Faculty, University of Zürich, Zürich, Switzerland

## Abstract

Accurate and rapid diagnostic tools are needed for management of the ongoing coronavirus disease 2019 (COVID-19) pandemic. Antibody tests enable detection of individuals past the initial phase of infection and will help to examine possible vaccine responses. The major targets of human antibody response in severe acute respiratory syndrome coronavirus 2 (SARS-CoV-2) are the spike glycoprotein (S) and nucleocapsid protein (N). We have developed a rapid homogenous approach for antibody detection termed LFRET (protein L-based time-resolved Förster resonance energy transfer immunoassay). In LFRET, fluorophore-labeled protein L and antigen are brought to close proximity by antigen-specific patient immunoglobulins of any isotype, resulting in TR-FRET signal generation.

We set up LFRET assays for antibodies against S and N and evaluated their diagnostic performance using a panel of 77 serum/plasma samples from 44 individuals with COVID-19 and 52 negative controls. Moreover, using a previously described S construct and a novel N construct, we set up enzyme linked immunosorbent assays (ELISAs) for antibodies against SARS-CoV-2 S and N. We then compared the LFRET assays with these enzyme immunoassays and with a SARS-CoV-2 microneutralization test (MNT).

We found the LFRET assays to parallel ELISAs in sensitivity (90-95% vs. 90-100%) and specificity (100% vs. 94-100%). In identifying individuals with or without a detectable neutralizing antibody response, LFRET outperformed ELISA in specificity (91-96% vs. 82-87%), while demonstrating an equal sensitivity (98%).

In conclusion, this study demonstrates the applicability of LFRET, a 10-minute ‘mix and read’ assay, to detection of SARS-CoV-2 antibodies.

## Introduction

In October 2020, the number of confirmed cases in the ongoing coronavirus disease 2019 (COVID-19) pandemic, caused by the severe acute respiratory syndrome coronavirus 2 (SARS-CoV-2), exceeded 40 million, with over a million deaths^1^. Reliable diagnostic assays are needed for specific management of COVID-19 patients as well as for epidemic surveillance and containment (“Test, trace and isolate”). Nucleic acid tests (NAT) or antigen tests serve to detect acute SARS-CoV-2 infection, whereas antibody testing tells the past-infection and/or immunity status, at both individual and population levels. Hence, antibody tests can be used for determining seroprevalences, examining vaccine responses in study settings, or determining whether an individual needs a booster as in the case of e.g. hepatitis B vaccine. With COVID-19, antibody testing may be the key in reaching the diagnosis for a patient presenting when the viral RNA has already waned, e.g. with late thromboembolic complications or prolonged symptoms^2^. The most widespread methods in antibody detection are enzyme immunoassays (EIAs) and lateral flow assays (LFAs); the former tend to be highly specific and sensitive yet require dedicated infrastructure and labor, and deliver the results at best within hours, whereas LFAs are simple and rapid but may be of substandard diagnostic performance.

We have previously set up rapid homogeneous (wash-free) immunoassays utilizing time-resolved Förster resonance energy transfer (TR-FRET)^3-8^. For FRET to occur, two fluorophores, donor and acceptor, are brought to close proximity, allowing excitation of the former to result in energy transfer to the latter, which then emits photons at a distinct wavelength. The efficiency of FRET is inversely dependent on the distance between the two fluorophores, with a 50 % efficiency typically achieved at 15 to 60 Å. To reduce background from the notoriously autofluorescent biological samples, a chelated lanthanide donor exhibiting long-lived fluorescence is employed, allowing for time-resolved measurement (TR-FRET). We have developed a rapid homogeneous TR-FRET-based immunoassay concept termed LFRET (protein L-based time-resolved Förster resonance energy transfer immunoassay) and demonstrated its excellent diagnostic performance in detection of antibodies against *Puumala orthohantavirus* nucleocapsid protein, *Zika virus* NS1 and the autoantigen tissue transglutaminase (tTG)^5,7,8^. LFRET relies on simultaneous binding to the antibody of interest of its donor-labeled antigen and of an acceptor-labeled protein L. If the patient’s serum contains antibodies against the antigen, they bring the two fluorophores to close proximity, generating a TR-FRET signal that indicates the presence of the specific antibodies. Interestingly, a recent report describes a TR-FRET based 1-hour assay for separate detection of anti-SARS-CoV-2 antibodies of diffent immunoglobulin isotypes^9^.

SARS-CoV-2 is an enveloped (+)ssRNA virus with a non-segmented 30 kb genome and four structural proteins: spike (S), envelope (E), membrane (M), and nucleocapsid (N). Protruding from the viral surface are transmembrane homotrimers of S, essential for host cell entry. The glycoprotein S is proteolytically cleaved into subunits S1 and S2, of which S1 contains the host cell receptor-binding domain (RBD), while S2 mediates fusion with the host cell membranes^10^. Like S, the E and M proteins are located on the viral envelope, whereas N protein binds the viral RNA to form a ribonucleoprotein complex that is encapsulated within the viral membrane.

Antibody responses to SARS-CoV-2 predominantly target the S and N proteins. In hospitalized patients, the median time from onset of symptoms to IgA, IgM and IgG seroconversion has been observed to be 11-14 days, with almost all individuals seroconverting by day 21^11-13^. The antibody levels correlate with the severity of disease, with few patients apparently not seroconverting^12^. Moreover, a fraction of the seroconverters do not seem to generate detectable neutralizing antibodies^14^. The neutralizing antibody (NAb) response correlates with the presence of anti-S antibodies^15,16^, with most but not all NAbs targeting the RBD^17^. IgG levels to other human betacoronaviruses have been observed to peak within months of infection and to wane within some years thereafter^18,19^. Moreover, reinfections with seasonal human coronaviruses have been observed as early as 12 months from the previous infection^20^. As for SARS-CoV-2, the persistence of antibodies and the extent to which these provide protective immunity remains as of yet uncertain.

In this study we introduce rapid wash-free LFRET assays for detection of antibodies against SARS-CoV-2 N and S antigens and compare them with ELISAs and microneutralization.

## Results

### LFRET incubation time, cutoff values and performance

LFRET assays for SARS-CoV-2 S and N were set up using Eu-labeled in-house antigens and AF-labeled protein L. First, the assay conditions were optimized separately for S and N using three known anti-S/-N ELISA-positive and three known anti-S/-N ELISA-negative samples (included in the full 129-sample panel). Thereafter the remaining 123 samples were tested in the optimized conditions. For detection of both anti-S and anti-N antibodies measurement at 7 minutes was found optimal.

Cutoffs for both anti-S and anti-N LFRET were set by measuring LFRET signals relative to buffer in 48 samples tested negative by anti-S and anti-N ELISA. The average plus four standard deviations (SD) was set as cutoff: 228.37 + 4 × 27.59 = 338.76 counts for anti-S and 220.94 + 4 × 27.73 = 331.86 counts for anti-N LFRET.

Performances of the anti-S and anti-N LFRET assays were then determined with the 129 samples including 77 sera or heparin/EDTA plasmas from 44 individuals with a previous RT-PCR-confirmed SARS-CoV-2 infection, four samples from four individuals negative for SARS-CoV-2 by both RT-PCR and serology, and 48 samples from individuals with a comprehensively negative SARS-CoV-2 serology. The sensitivities and specificities of SARS-CoV-2 anti-S and anti-N LFRET in detection of PCR-positive individuals were 87% and 100% & 78% and 100%, respectively. The combined anti-S/-N LFRET sensitivity and specificity were 90% and 100%: if either anti-S or -N LFRET was positive, the composite result was considered positive (Table 1). The development of LFRET signals over time among patients with follow-up samples available is shown in Figure S1.

**Table 1.**
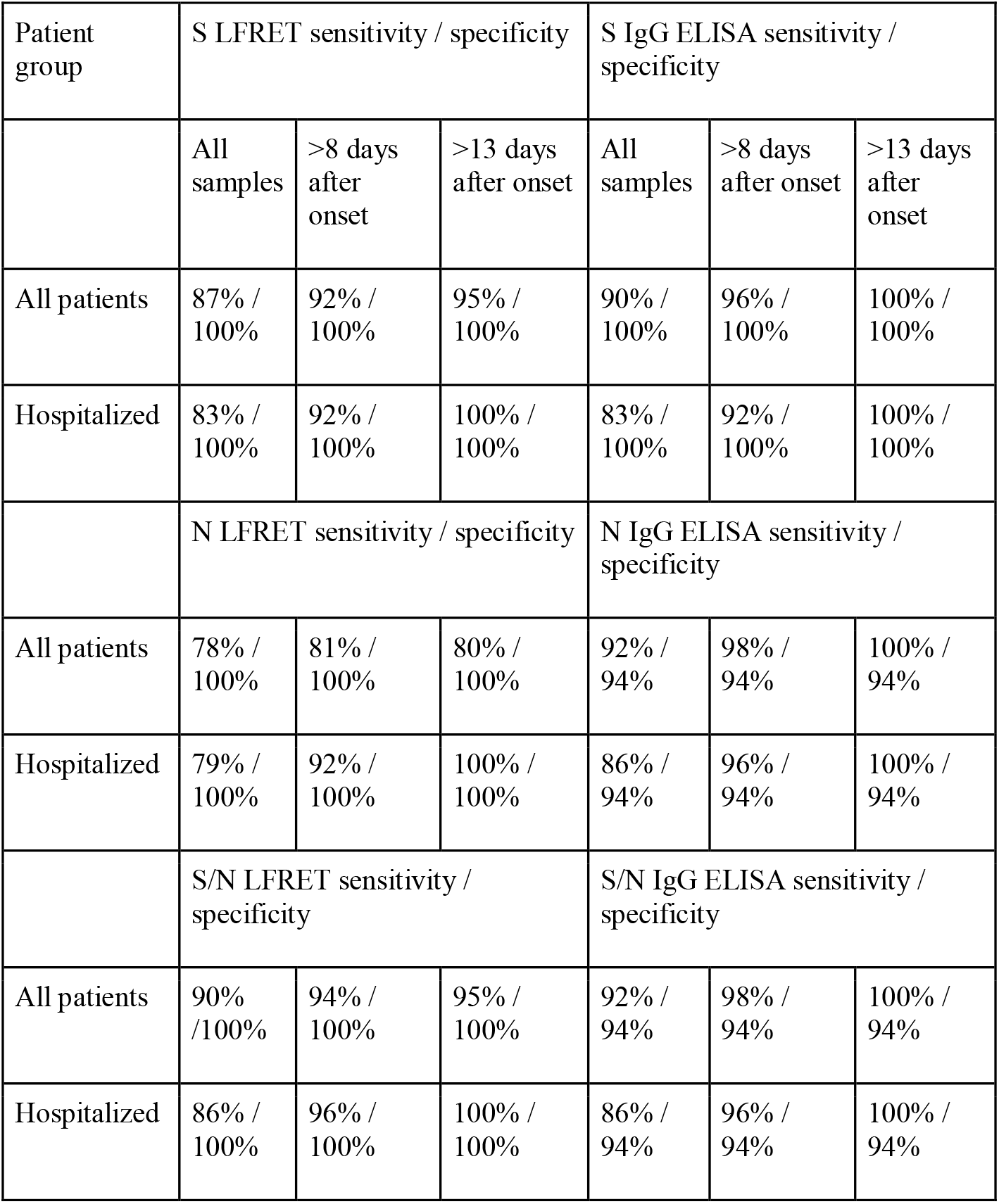
Sensitivity and specificity of ELISA and LFRET in detection of SARS-CoV-2-PCR-positive individuals for all patients and hospitalized patients at different time points from symptom onset. Altogether twenty samples from ten PCR-positive individuals for whom the onset of symptoms is unknown are excluded. Specificity is calculated from non-hospitalized asymptomatic individuals. N = nucleocapsid protein. S = spike glycoprotein. LFRET = protein L–based time-resolved Förster resonance energy transfer immunoassay. ELISA = enzyme-linked immunosorbent assay.

| Patient group | S LFRET sensitivity / specificity |  |  | S IgG ELISA sensitivity / specificity |  |  |
| --- | --- | --- | --- | --- | --- | --- |
|  | All samples | >8 days after onset | >13 days after onset | All samples | >8 days after onset | >13 days after onset |
| All patients | 87% / 100% | 92% / 100% | 95% / 100% | 90% / 100% | 96% / 100% | 100% / 100% |
| Hospitalized | 83% / 100% | 92% / 100% | 100% / 100% | 83% / 100% | 92% / 100% | 100% / 100% |
|  | N LFRET sensitivity / specificity |  |  | N IgG ELISA sensitivity / specificity |  |  |
| All patients | 78% / 100% | 81% / 100% | 80% / 100% | 92% / 94% | 98% / 94% | 100% / 94% |
| Hospitalized | 79% / 100% | 92% / 100% | 100% / 100% | 86% / 94% | 96% / 94% | 100% / 94% |
|  | S/N LFRET sensitivity / specificity |  |  | S/N IgG ELISA sensitivity / specificity |  |  |
| All patients | 90% / 100% | 94% / 100% | 95% / 100% | 92% / 94% | 98% / 94% | 100% / 94% |
| Hospitalized | 86% / 100% | 96% / 100% | 100% / 100% | 86% / 94% | 96% / 94% | 100% / 94% |

### ELISAs and microneutralization

In order to compare the performance of LFRET with classical serology, we tested the set of samples described above with SARS-CoV-2 anti-S and anti-N IgA, IgM and IgG ELISAs as well as SARS-CoV-2 microneutralization. The panel of seronegatives was excluded from anti-N IgA and IgM as well as anti-S IgM ELISAs. Altogether 107 samples underwent microneutralization, including 64 samples from RT-PCR positive patients and 43 seronegative samples. Microneutralization titers of ≥20 were considered positive.

The ELISA cutoffs were set at average plus four standard deviations of absorbances measured from 14 serum samples from SARS-CoV-2 seronegative Department staff members.

The sensitivities and specificities of ELISA for anti-S IgA, IgG and IgM in samples from SARS-CoV-2 RT-PCR-positive individuals were 91% and 98%, 90% and 100% & 66% and 100%, respectively. The corresponding sensitivities and specificities of anti-N ELISA were 75% and 100% (IgA), 92% and 94% (IgG) & 16% and 100% (IgM), respectively (Figure S2). Pearson correlation between anti-S and anti-N IgG ELISAs was 0.90, 0.79 between IgM ELISAs and 0.31 between IgA ELISAs.

### Comparison of LFRET, ELISA and microneutralization

Comparison between the LFRET signals and ELISA absorbances is presented in Figure 2. For anti-N antibodies, the correlation between LFRET and IgA or IgM ELISA results was low (R=0.25 for IgA and R=0.13 for IgM). With IgG ELISA a stronger correlation of R=0.62 was seen, apparently hampered by saturation of the ELISA signal. For anti-S-antibodies, correlations between IgA, IgG and IgM ELISAs were R=0.52, R=0.62 and R=0.56, respectively. Higher LFRET signals were seen in samples from patients with severe disease, especially in anti-S LFRET and when samples taken less than two weeks from onset were excluded (Figure S3). The agreement between anti-N ELISA and LFRET was 88-89%, and that between anti-S ELISA and LFRET 96-98% (Table S1). The samples representing discordance between PCR, LFRET and/or ELISA are detailed in Table S2.

**Figure 1.**
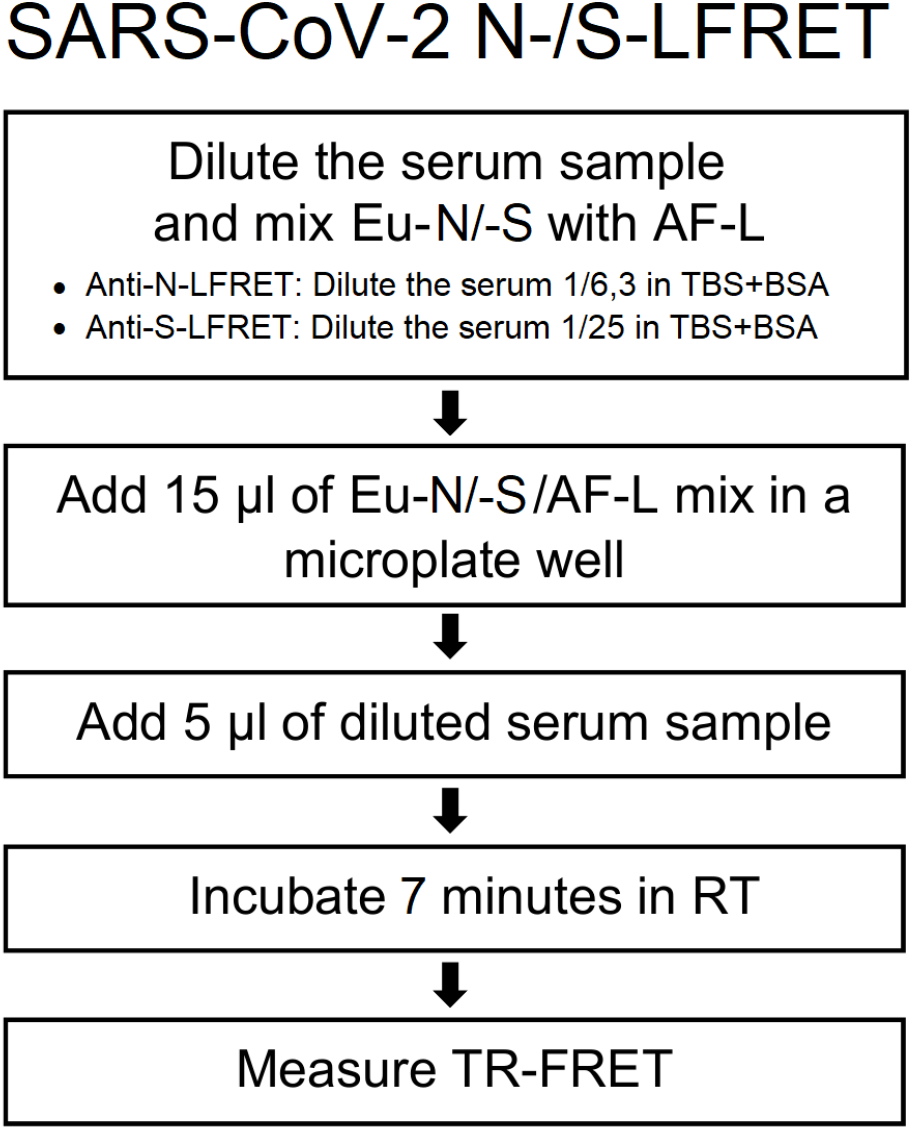
Simplified protocol for SARS-CoV-2 N and S LFRET assay. Eu-N/-S = Europium-labeled nucleocapsid protein/spike glycoprotein. AF-L = Alexa Fluor™ 647 - labeled protein L. TR-FRET = time-resolved Förster resonance energy transfer. RT = room temperature. TBS+BSA (50mM Tris-HCl, 150mM NaCl, pH 7.4, 0.2% BSA) was used for all dilutions. On-plate dilutions were 5 nM Eu-N/500 nM AF-L/serum 1/25 for anti-N and 5 nM Eu-S/250 nM AF-L/serum 1/100 for anti-S LFRET. For further details see the prior publication^5^.

**Figure 2.**
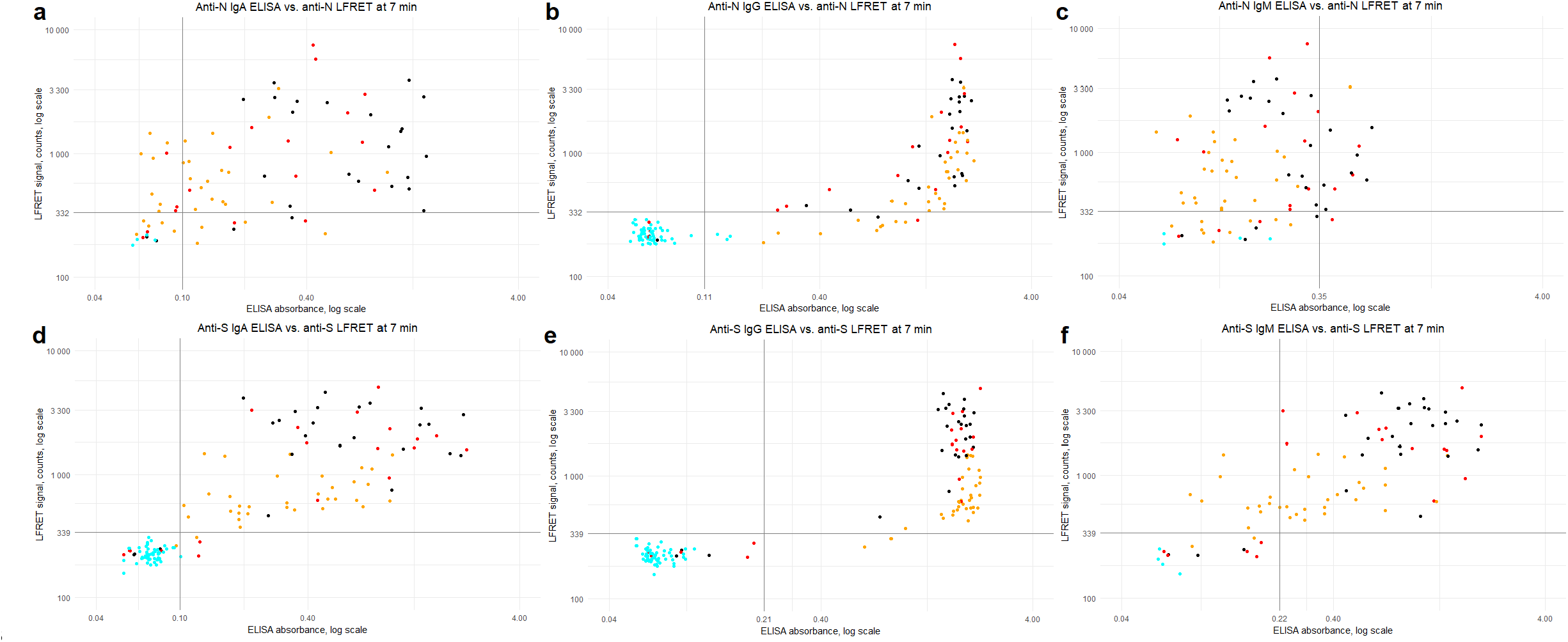
ELISA (x-axis) vs. LFRET (y-axis) results by disease severity. a) Anti-N IgA ELISA vs. anti-N LFRET (N=81, R=0.25). b) anti-N IgG ELISA vs. anti-N LFRET (N=129, R=0.62). c) anti-N IgM ELISA vs. anti-N LFRET (N=81, R=0.13). d) anti-S IgA ELISA vs. anti-S LFRET (N=129, R=0.53). e) anti-S-IgG ELISA vs. anti-S LFRET (N=129, R=0.62). f) anti-S IgM ELISA vs. anti-S LFRET (N=81, R=0.56). Colour of the dot indicates SARS-CoV-2 PCR result and disease severity: cyan = PCR negative; yellow = non-hospitalized, PCR-positive; red = non-ICU hospitalized, PCR positive; black = hospitalized in ICU, PCR positive. Horizontal and vertical black lines indicate LFRET and ELISA cutoffs. On the X-axis, ELISA absorbance on a logarithmic scale and on the y-axis, LFRET signal on a logarithmic scale. S = spike glycoprotein. N = nucleocapsid protein. LFRET = protein L–based time-resolved Förster resonance energy transfer immunoassay. ELISA = enzyme immunoassay. R = Pearson’s correlation coefficient.

The LFRET and ELISA results are compared with microneutralization titers in Figure 3. Spearman’s rank correlation coefficient (rho) between anti-S LFRET and MNT was 0.87, whereas for anti-S IgG, IgA and IgM ELISAs it was 0.68, 0.86 and 0.81, respectively. Likewise, between anti-N LFRET and MNT Spearman’s rho was 0.83, while 0.81, 0.69 and 0.61 for anti-N IgG, IgA and IgM ELISAs. Higher neutralization titers were observed in samples from hospitalized individuals. Sensitivities and specificities of LFRET and ELISA in correctly identifying microneutralization-positive and -negative samples are shown in Table S3.

**Figure 3.**
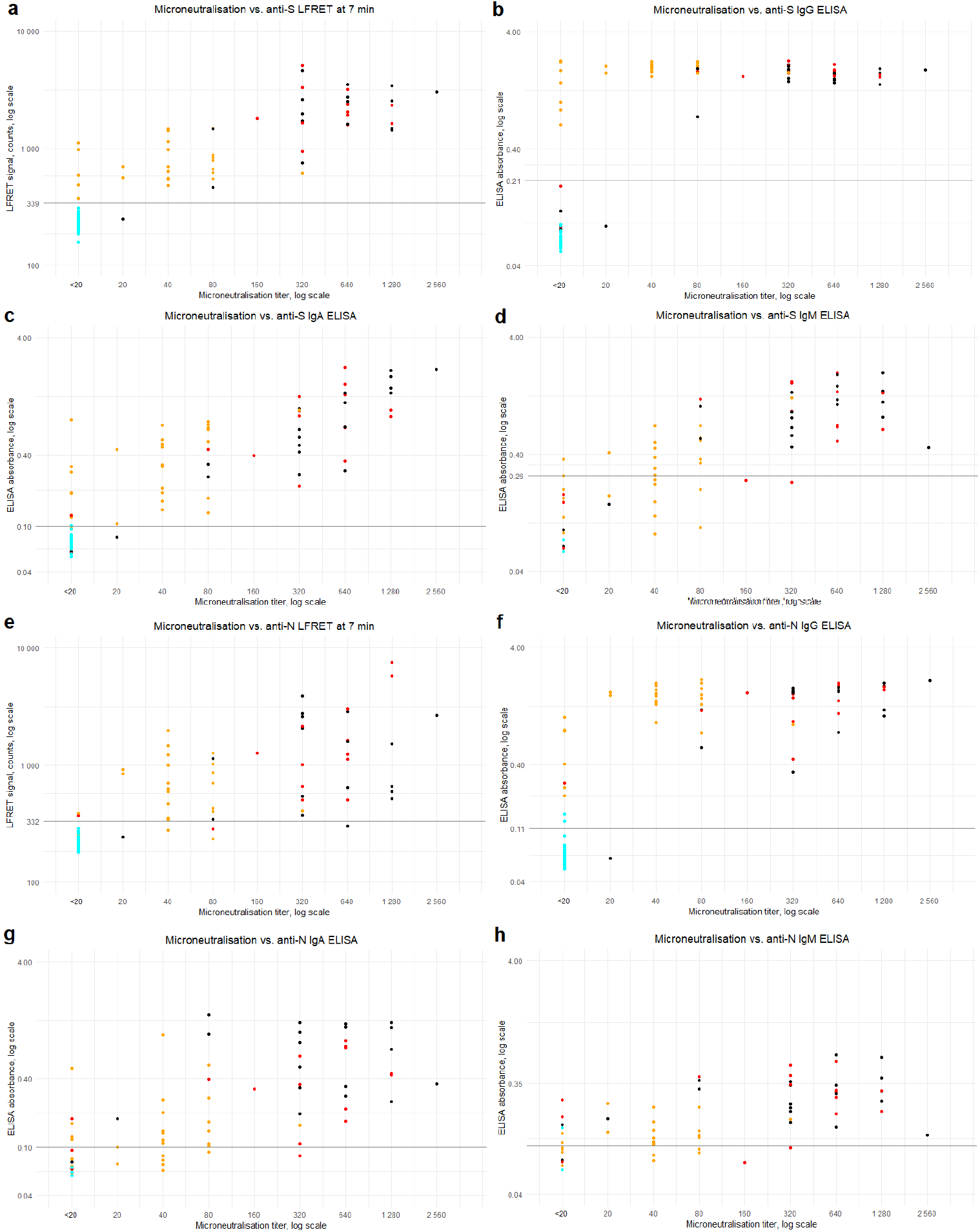
Microneutralization vs. LFRET and ELISA. Microneutralization titers are on the x-axis and LFRET signal or ELISA absorbance on the y-axis. Logarithmic scale is used on both axes. a) Microneutralization titer vs. anti-S LFRET signal (N=107, ρ=0.87). b-d) Microneutralization titer vs. anti-S IgG, IgA and IgM ELISA (N=107, 107 and 67, ρ=0.68, 0.86 and 0.81). e) Microneutralization titer vs. anti-N LFRET signal (N=107, ρ=0.83). f-h) Microneutralization titer vs. anti-N IgG, IgA and IgM ELISA (N=107, 67 and 67, ρ=0.81, 0.69 and 0.61). Colour of the dots indicate SARS-CoV-2 PCR result and disease severity: cyan = PCR negative; yellow = non-hospitalized, PCR-positive; red = non-ICU hospitalized, PCR positive; black = hospitalized in ICU, PCR positive. Horizontal black lines indicate LFRET/ELISA cutoffs. S = spike glycoprotein. N = nucleocapsid protein. LFRET = protein L–based time-resolved Förster resonance energy transfer immunoassay. ELISA = enzyme immunoassay. ρ = Spearman’s rank correlation coefficient.

### Receiver operating characteristic analysis

We also rated the performances of both LFRET assays using receiver operating characteristic (ROC) curves (Figure S4). The curves were plotted in RStudio (version 1.3.1073) with the ROCR library (version 1.0-11). With the assumption that a positive result in either IgM, IgG or IgA ELISA for a given sample signifies ‘true’ positivity, the respective areas under the curve (AUCs) for both anti-N and anti-S LFRET assays were very high, 0.94 and 0.97.

## Discussion

We set up rapid LFRET immunoassays for detection of anti-SARS-CoV-2 S and N antibodies for identification of individuals exhibiting an immune response against SARS-CoV-2. Management of both COVID-19 patients and the ongoing pandemic at the population level calls for accurate diagnostic tools applicable in various settings, including resource poor areas without central laboratory facilities. Antibody assays allow detection of individuals past the initial infection phase as well as assessment of a possible vaccine response.

We have previously applied LFRET to diagnostics of viral and autoimmune diseases. Here, we further reduced the incubation time from the prior 20-30 to 7 minutes without compromising sensitivity or specificity. The simple, rapid ‘mix and read’ workflow of the assay could allow faster turnaround time from sample arrival to results as well as higher throughput compared to the currently popular EIAs. Moreover, the ease and speed of performing LFRET makes it more feasible for use in diverse enviroments, including point-of-care and limited-resource settings.

Combined anti-S/-N LFRET (i.e. if either assay is positive, the composite result is positive) was equal to anti-S IgG ELISA in terms of sensitivity (90%) and specificity (100%) in identification of RT-PCR-positive individuals (Table 1). Considering that the median time from symptom onset to seroconversion is 1-2 weeks for SARS-CoV-2^11-13^, we can better assess the sensitivities of these assays by excluding samples taken before two weeks from symptom onset. Excluding these samples, the sensitivities of anti-S, anti-N and anti-S/-N LFRET are 95%, 80% and 95%, respectively. Simultaneously, the sensitivities of anti-S/-N IgG ELISAs increase to 97%, 100% and 100%. Combining sensitivity and specificity into a single value, the AUCs of anti-N and -S LFRET assays – 0.94 and 0.97 – reflect excellent performance (Figure S4).

Anti-SARS-CoV-2 antibody levels are higher in patients with severe clinical presentation^12,15^. Also in our study the LFRET signals of hospitalized patients exceeded those of non-hospitalized yet PCR-positive individuals (Figures 2 and S3). In the hospitalized COVID-19 patients, the sensitivity of both anti-S and anti-N LFRET increased to 100% by two weeks from symptom onset (Table 1), importantly for clinical use. In follow-up, the LFRET signals first showed a rapid rise within three weeks from onset and thereafter plateaued or slowly declined (Figure S1).

The agreement between ELISA and LFRET was high: ∼90% for anti-N-antibodies and >95% for anti-S-antibodies (Table S1). With the anti-N and anti-S LFRET vs. IgG ELISA results combined, the overall agreement between the methods in the entire study material was 94% (124/129 samples). A closer look at the discordance (Table S2) shows three samples (65, 72 and 86) from PCR-positive individuals who remained seronegative in both LFRET and ELISA, likely due to early sampling (Figure S1). Four samples (71, 70, 7 and 24) were negative in LFRET but positive in PCR and ELISA: The first two were early infection specimens (taken 8 and 13 days post onset of symptoms) positive in anti-N IgA ELISA, suggestive of early IgA seroconversion, a phenomenon observed previously^21^. The other two were taken 4 and 9 weeks after onset from non-hospitalized patients positive in anti-S and -N IgG and/or IgA ELISAs. These ELISA reactivities were weak (Figure 2e), suggesting that the negativity in LFRET might reflect lower analytical sensitivity. Two samples (82, 92) were negative in LFRET and RT-PCR but positive in anti-N IgG ELISA. Additionally, two samples (103, 121) were negative in LFRET and MNT, but positive in either anti-N IgG ELISA or anti-S IgA ELISA. No false positives were observed in LFRET.

In microneutralization, all but one of the reactive samples were also positive in anti-S LFRET, IgG and IgA ELISA and anti-N IgG ELISA (Figure 3), the exception being an ICU patient sampled 13 days after onset with an MNT titer of 20 and a positive anti-N IgA ELISA (Table S2, sample 70). Some of the studied individuals who seroconverted did not exhibit a detectable neutralizing antibody response, as observed previously^14^. Interestingly, the specificities of the LFRET assays in identification of the non-neutralizing individuals as negatives (91% for S and 96% for N) were higher than those of IgG ELISAs (87% for S and 82% for N) (Table S3). This may be due to lower analytical sensitivity of LFRET, as the undetectable neutralization could result from lower overall levels of anti-SARS-CoV-2 antibodies in the LFRET-negative but ELISA-positive samples. Nevertheless, among the assays evaluated, anti-S LFRET demonstrated the best overall performance in identification of samples containing neutralizing antibodies, with a sensitivity of 98% and a specificity of 91%.

Our study has some limitations. First, our SARS-CoV-2-positive samples originated from symptomatic patients. Individuals with asymptomatic infection may mount a significantly lower antibody response^22^, whereby the sensitivity of LFRET in such individuals might be lower. A second limitation is that antibodies against other coronaviruses, especially the widely circulating OC43, HKU1, NL63 and 229E, were not examined. Those antibodies potentially cross-reacting in the SARS-CoV-2 assay could reduce its specificity. However, the RT-PCR and neutralization results strongly indicate that the observed antibody responses were SARS-CoV-2-specific.

In conclusion, this study demonstrates the applicability of the LFRET approach to detection of SARS-CoV-2 antibodies. While in sensitivity and specificity the assay appears to parallel ELISA, the new assay is as easy and rapid to perform as an LFA, requiring only combination of the diluted sample with a reagent mix and reading the result after 7 minutes. In prediction of neutralization capacity, the anti-S LFRET outperformed ELISA in specificity, at equal sensitivity.

## Materials and Methods

### Samples

This study included 77 serum/plasma samples from 40 individuals tested positive and four samples from four individuals tested negative for SARS-CoV-2 by RT-PCR from nasopharyngeal swab samples. The positive samples were taken at 8 to 81 days after onset of symptoms. Additionally, 48 serum samples from asymptomatic individuals with a comprehensively negative SARS-CoV-2 serology (Euroimmun IgG, IFA IgG virus, IFA IgG spike, microneutralization negative) were included in the study. The data and samples were collected under research permit HUS/211/2020 and ethics committee approval HUS/853/2020 (Helsinki University Hospital, Finland).

### Nucleic acid testing

Nucleic acid testing for SARS-CoV-2 was done from nasopharyngeal swab samples with either the Cobas® SARS-CoV-2 test using the Cobas® 6800 system (Roche Diagnostics, Basel, Switzerland), a protocol based on one previously described by Corman et al.^23^, or the Amplidiag® COVID-19 test (Mobidiag, Espoo, Finland).

### Molecular cloning

For protein expression, we acquired the ZeoCassette Vector (pCMV/Zeo) from ThermoFisher Scientific, and excised the Zeocin resistance gene from the vector using FastDigest EcoRI and XhoI (ThermoFisher Scientific) according to manufacturer’s protocol. The excised gene was agarose gel purified, blunted using T4 DNA polymerase (ThermoFisher Scientific), and purified using Ampure XP beads (Beckman Coulter) both following the manufacturer’s protocol. The selection gene was inserted into pCAGGS/MCS and to the pCAGGS vector bearing SARS-CoV-2 S protein (described in ^24,25^) gene by treating the plasmids with FastDigest SapI/LguI (ThermoFisher Scientific) according to manufacturer’s protocol, followed by blunting and purifications as above. The insert was ligated to the plasmids using T4 DNA ligase (ThermoFisher Scientific) according to manufacturer’s protocol, the ligation products transformed into *Escherichia coli* (DH5a strain), followed by plating the bacteria onto LB plates with 100 µg/ml of ampicillin and 50 µg/ml Zeocin (ThermoFisher Scientific). After overnight incubation at 37 °C, single colonies were picked and grown in 5 ml of 2xYT medium supplemented with 100 µg/ml of ampicillin and 50 µg/ml Zeocin overnight at 37 °C. The plasmids were purified using GeneJET Plasmid Miniprep Kit (ThermoFisher Scientific), and plasmids bearing the insert identified by restriction digestion (FastDigest EcoRI, ThermoFisher Scientific) and agarose gel electrophoresis. For both constructs, clones with the insert in reverse and forward direction were selected for ZymoPURE II Plasmid Maxiprep Kit (ZymoResearch) preparations done following manufacturer’s guidelines. A synthetic SARS-CoV-2 NP gene under Kozak sequence and a signal sequence MMRPIVLVLLFATSALA flanked by KpnI and SgsI restriction sites was obtained from ThermoFisher Scientific. The SARS-CoV-2 NP cassette was subcloned into pCAGGS/MCS-Zeo-fwd vector and plasmid maxipreps prepared as described above.

### Protein expression and purification

We initially attempted producing SARS-CoV-2 S protein in Expi293F cells utilizing the Expi293 Expression System (ThermoFisher Scientific). Briefly, the pCAGGS plasmid bearing codon-optimized SARS-CoV-2 spike^24^ was transiently transfected into Expi293F cells as advised^24,25^, except that we used spinner flasks (disposable 125 ml spinner flask, Corning) for the culture. Protein purification from supernatants collected at five days post transfection followed the protocol described^25^ and yielded 0.2-0.3 mg per 100 ml culture, in line with earlier reports^24,25^.

Next, we transfected adherent HEK293T cells with pCAGGS-SARS-CoV-S-Zeo plasmids using Fugene HD at 3.5:1 ratio, in suspension as described^26^. The transfected cells were plated onto six-well plates, and at 48 h post transfection subjected to Zeocin selection, 150 µg/ml Zeocin in high glucose DMEM (Dulbecco’s Modified Eagle’s Medium, Sigma) supplemented with 5% fetal bovine serum (Gibco) and 4 mM L-glutamine. Two days after initiating the selection, the cells were trypsinized and transferred into fresh wells, with fresh media and antibiotics provided at 2 to 3 day intervals. Once confluent, the cells were trypsinized, counted (TC20 cell counter, Bio-Rad), diluted to ∼30 cells/ml, and dispensed onto 96-well plates, 100 µl per well. After confluency, we switched to serum-free FreeStyle 293 Expression Medium (ThermoFisher Scientific) with 100 µg/ml Zeocin, and incubated the cells at 37 °C 5% CO2. At 48 h, we analyzed the medium for the presence of SARS-CoV-2 S protein by dot blotting, briefly 2.5 µl of the supernatant was dried onto a nitrocellulose membrane, the membrane blocked (3% skim milk in Tris-buffered saline with 0.05% Tween-20), washed, probed with rabbit anti-RBD (Sino Biological, 40592-T62), washed, probed with anti-rabbit IRDye800 (LI-COR Biosciences), washed, and read using Odyssey Infrared Imaging System (LI-COR Biosciences). The clone with the highest amount of SARS-CoV-2 S protein in the cell culture supernatant, HEK293T-spike-D5, was then expanded in DMEM with 5% FBS, 4 mM L-glutamine and 100 µg/ml Zeocin, and ampouled for storage in liquid nitrogen. We adapted the HEK293T-spike-D5 cells for suspension culture by placing trypsinized cells into a spinner flask with Expi293 Expression Medium (ThermoFisher Scientific) with 100 µg/ml of Zeocin. We stored an aliquot of the adapted cells in liquid nitrogen and tested their ability to produce SARS-CoV-2 S protein in both Expi293 and FreeStyle 293 Expression Medium (both ThermoFisher Scientific). Having been cultured in the spinner flask for 5 to 8 days, the cells reached a density of >3 × 10^6^ cells/ml. The protein from the supernatants was purified using the protocol described^25^, with yields of 0.8-1.2 mg per 100 ml culture.

For production of SARS-CoV-2 N protein, we transfected Expi293F cells with pCAGGS-SARS-CoV-2-NP-Zeo using Fugene HD, briefly, 100 µg of the plasmid diluted into 10 ml of OptiMEM (Sigma), 350 µl of Fugene HD added followed by mixing and 15 min incubation at room temperature, after which the plasmid mix was added onto Expi293F cells in Expi293 Expression Medium at 2.5 × 10^6^ cells/ml. The incubation of the transfected cells was continued for four days, after which the supernatant was collected, and the protein purified (yield approximately 1 mg per 100 ml culture) as described for S protein^25^. The remaining cells were treated with 0.25% Trypsin-EDTA (Sigma) to remove dead cells, and after two washes were put back into the spinner flask with fresh Expi293 Expression Medium supplemented with 100 µg/ml of Zeocin. Eventually a population of cells started to proliferate (Expi-NP-zeo cells), and were aliquoted in liquid nitrogen. The protein production and purification occurred as described above with yields of ∼1 mg per 100 ml culture.

### Protein labeling

We labeled the SARS-CoV-2 S and N proteins with the donor fluorophore europium (Eu) using QuickAllAssay Eu-chelated protein labeling kit (BN Products and Services Oy) according to the manufacturer’s instructions to generate Eu-labeled S (Eu-S) and N (Eu-N). We also labeled recombinant protein L (Thermo Scientific) with the acceptor fluorophore Alexa Fluor 647 to generate AF647-labeled protein L (AF-L), as reported^5^. IgG-free bovine serum albumin (BSA) was from Jackson ImmunoResearch Inc.

### TR-FRET assays

The LFRET assay was done as described^5^ and as briefly illustrated by the flowchart in Figure 1. For calculating the relative TR-FRET signal increase, we here replace the pool of negative sera with TBS-BSA (50mM Tris-HCl, 150mM NaCl, pH 7.4, 0.2% BSA). To establish LFRET assays for S and N, we optimized the component concentrations by cross-titration using three positive and three negative serum samples. For detection of anti-N antibodies, we found the optimal on-plate dilution for serum to be 1/25, and the optimal on-plate concentrations for AF-L and Eu-N to be 500 nM and 5 nM, respectively. For detection of anti-S antibodies, we found an on-plate dilution of 1/100 for serum, AF-L concentration of 250 nM and Eu-S concentration of 5 nM optimal. We measured TR-FRET at 0, 7, 15, 22, 30, 45 and 60 minutes after combining the reagents. TR-FRET counts were measured with Wallac Victor^2^ fluorometer (PerkinElmer) and normalized as described^3^.

### Enzyme linked immunosorbent assays (ELISAs)

We set up the SARS-CoV-2 S protein ELISA as described^25^ with the following amendments. We coated the plates (ThermoScientific NUNC-immuno 446442 polysorp lockwell C8) with 50 ul/well of antigens diluted 1 µg/ml into 50 mM carbonate-bicarbonate buffer pH 9.6 (Medicago AB) and used 1-Step Ultra TMB-ELISA Substrate Solution (ThermoFisher Scientific). As secondary antibodies we made use of polyclonal rabbit anti-human IgA-horeseradish peroxidase (HRP), -IgM-HRP, and -IgG-HRP (all from Dako) at respective dilutions of 1:5000, 1:1500, and 1:6000. The colorimetric reaction was terminated by addition to 0.5 M sulphuric acid (Fluka), and the absorbances recorded (HIDEX Sense) at 450 nm. The N protein ELISA followed the same protocol.

### Microneutralization

For the SARS-CoV-2 microneutralization assay we first cultured Vero E6 cells on 96-well plates (Thermo Scientific) overnight at +37°C in 2% MEM (Eagle Minimum Essential Media [Sigma-Aldrich] supplemented with 2% inactivated fetal bovine serum [Thermo Scientific], 2 mM L-glutamine [Thermo Scientific], 100 units penicillin, and 100 µg/ml streptomycin [Sigma-Aldrich]). The following day we made a two-fold dilution series (1:20 to 1:1280) of the serum samples in 2% MEM and combined 50 µl of each dilution with 50 µl of virus (1000 plaque forming units (pfu)/ml in 2% MEM). The serum-virus mixes were kept for 1h at +37°C. The cells were inoculated with the serum-virus mixes and grown in incubators at +37°C. After 4 days, the cultures were fixed with formalin, stained with crystal violet and the neutralization titers recorded.

## Data Availability

All study data are included in the article and the supplementary information.

## Acknowledgments and funding information

The authors wish to acknowledge Drs. Tomas Strandin and Eliisa Kekäläinen for helping in sample collection. Reagent generation in the Krammer laboratory was supported by the NIAID Centers of Excellence for Influenza Research and Surveillance (CEIRS) contract HHSN272201400008C and the Collaborative Influenza Vaccine Innovation Centers (CIVIC) contract 75N93019C00051.

The authors received funding for this work from the following sources: the Sigrid Jusélius Foundation, the Magnus Ehrnrooth Foundation, Finnish Society of Sciences and Letters, the Research Funds of University of Helsinki and Helsinki University Hospital (TYH 2018322), Finska Läkaresällskapet, the Finnish Medical Foundation, Academy of Finland (#1308613, #1336490, #336439 and #335527), Juho Vainio Foundation, Jane and Aatos Erkko Foundation, the European Union Horizon 2020 programme VEO (Versatile emerging infectious disease observatory, grant No. 874735) and from Private donors through the University of Helsinki.

