## Supplementary information for "Rapid homogeneous assay for detecting antibodies against SARS-CoV-2"

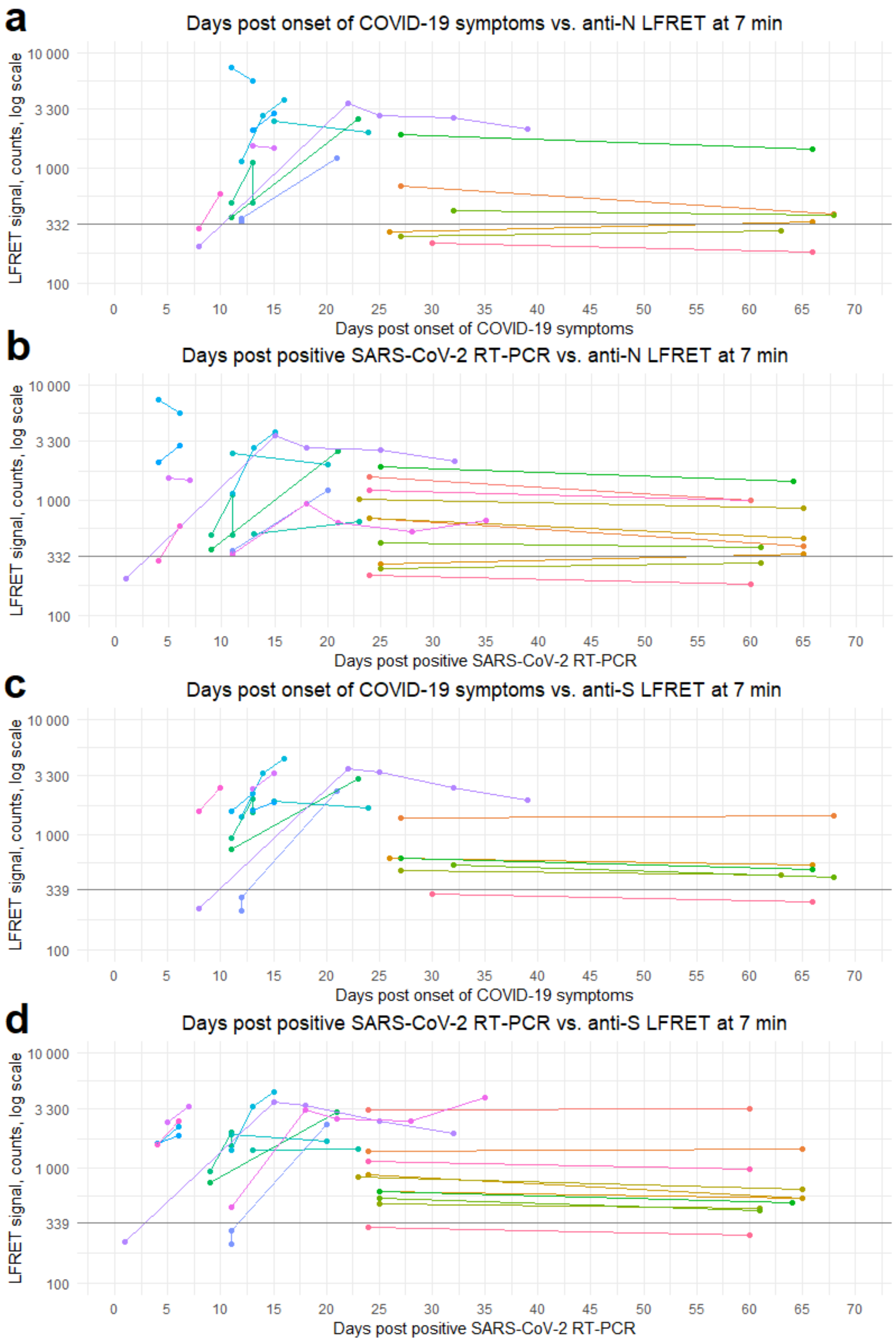


**Figure S1**. Development of LFRET signals in follow-up samples. a) Days post onset of COVID-19 symptoms (x-axis) vs anti-N LFRET signal (y-axis) (N=38 samples from 16 individuals). b) Days post positive SARS-CoV-2 PCR (x-axis) vs anti-N LFRET signal (y-axis) (N=53 samples from 22 individuals). c) Days post onset of COVID-19 symptoms (x-axis) vs anti-S LFRET signal (y-axis) (N=38 samples from 16 individuals). d) Days post positive SARS-CoV-2 PCR (x-axis) vs anti-S LFRET signal (y-axis) (N=53 samples from 22 individuals). Horizontal line is the LFRET cutoff. S = spike glycoprotein. N = nucleocapsid protein. LFRET = protein L–based time-resolved Förster resonance energy transfer immunoassay.

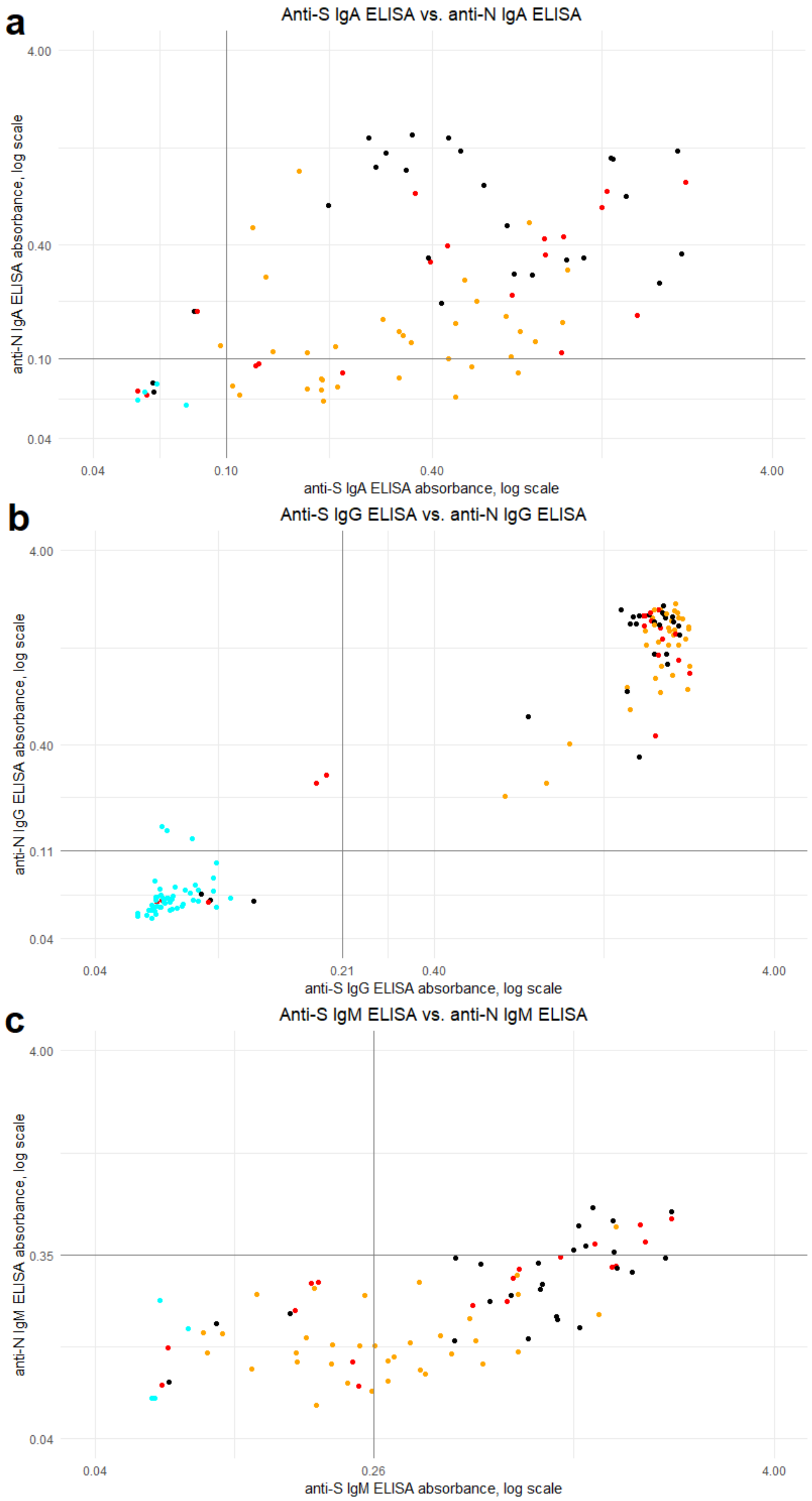


**Figure S2.** SARS-CoV-2 spike glycoprotein (S) versus nucleocapsid protein (N) ELISA. a) anti-S vs. anti-N IgA ELISA results (N=81, R=0.31) b) anti-S vs. anti-N IgG ELISA results (N=129, R=0.90). c) anti-S vs. anti-N IgM ELISA results (N=81, R=0.79). On the x-axis, anti-S ELISA absorbance, on the y-axis anti-N ELISA absorbance. Colour of the dot indicates SARS-CoV-2 PCR result and disease severity: cyan = PCR negative; yellow = non-hospitalized, PCR-positive; red = non-ICU hospitalized, PCR positive; black = hospitalized in ICU, PCR positive. Horizontal and vertical lines indicate ELISA cutoffs. R = Pearson’s correlation coefficient.

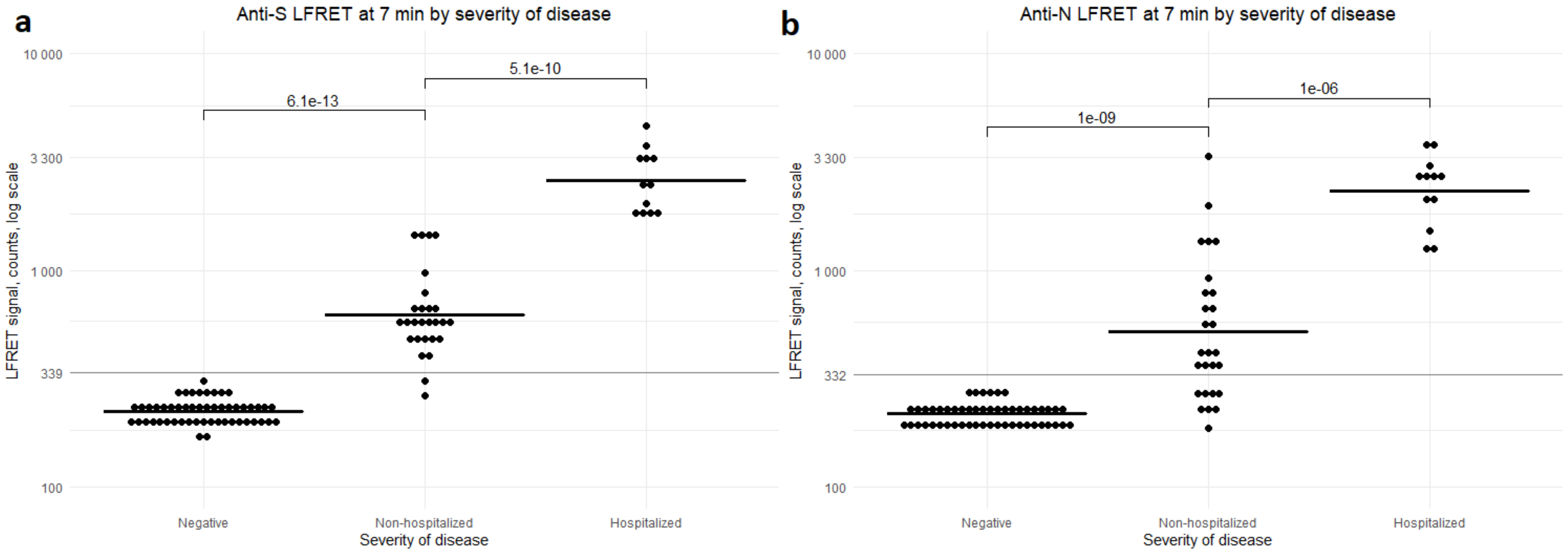


**Figure S3.** LFRET vs. severity of disease in samples taken after two weeks from onset of disease. a) Severity of disease (x-axis) vs. anti-S LFRET (y-axis) b) Severity of disease (x-axis) vs. anti-N LFRET (y-axis). Negative = samples from comprehensively SARS-CoV-2 seronegative individuals (N=52). Non-hospitalized = samples from non-hospitalized COVID-19 patients (N=27). Samples from hospitalized individuals with COVID-19 (N=12) are grouped together to include samples from individuals treated outside of ICU (N=3) and in ICU (N=9). Horizontal lines indicate LFRET cutoffs (thin grey lines) and mean LFRET signal (thick black lines). Logarithmic scale is used on the y-axis. Differences between groups are statistically significant, as indicated by the p values within the figure. S = spike glycoprotein. N = nucleocapsid protein. LFRET = protein L–based time-resolved Förster resonance energy transfer immunoassay.

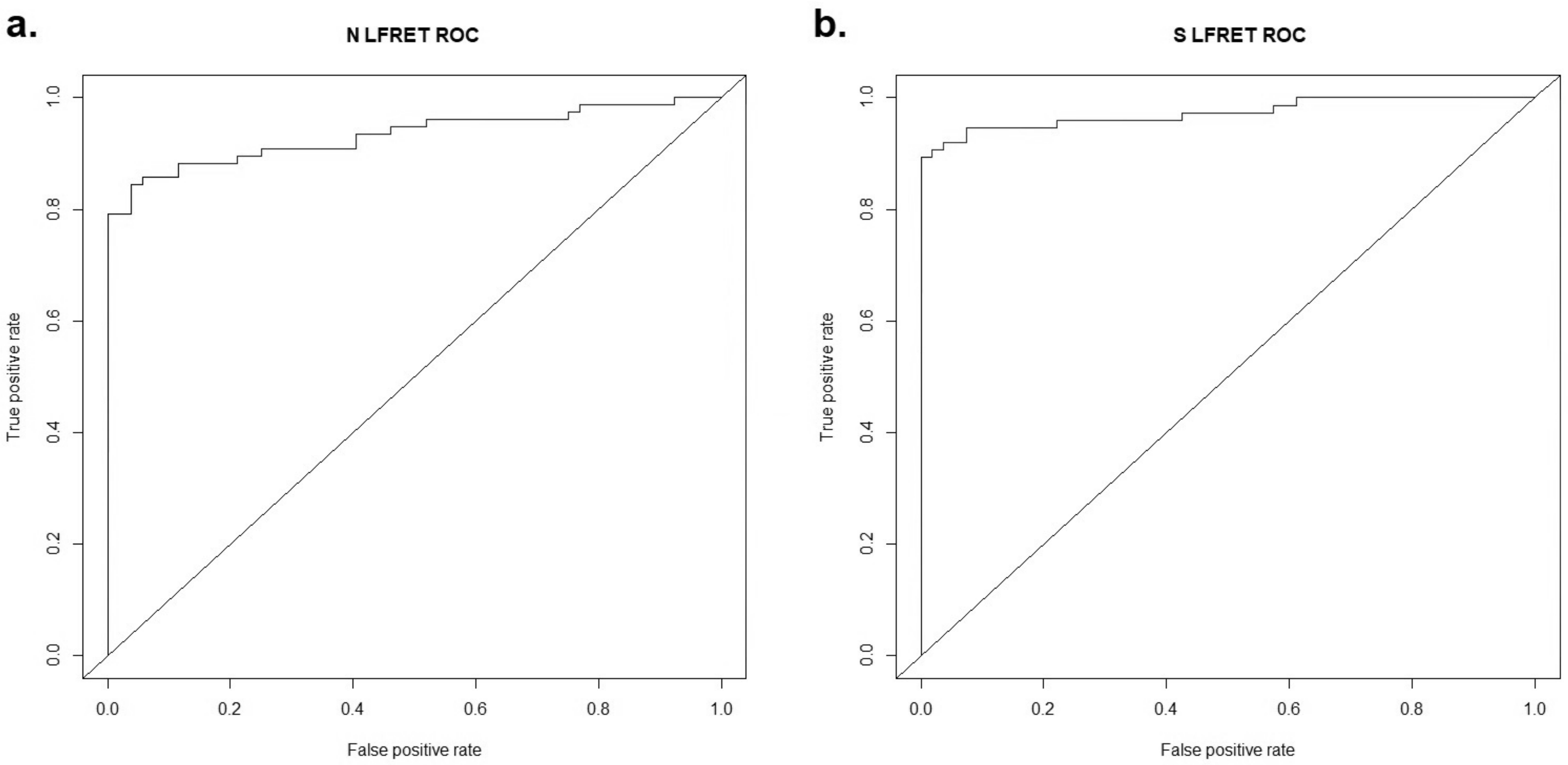


**Figure S4.** Receiver operating character (ROC) curves for LFRET. a) anti-N LFRET. AUC = 0.94. b) anti-S LFRET. AUC= 0.97. False positive rate on the x-axis, true positive rate on the y-axis. Enzyme immunoassay (IgA, IgG and IgM results combined) is used as the reference. N = nucleocapsid protein. S = spike glycoprotein. LFRET = protein L–based time-resolved Förster resonance energy transfer immunoassay.

**Table S1**. Agreement between ELISA and LFRET. Ig ELISA = IgA, IgG and IgM results combined: if any of these are positive, total Ig is considered positive. N/S ELISA/LFRET: if either N or S ELISA/LFRET is positive, the N/S result is considered positive. Number of samples in brackets. S = spike glycoprotein. N = nucleocapsid protein. LFRET = protein L–based time-resolved Förster resonance energy transfer immunoassay. ELISA = enzyme immunoassay. MNT = microneutralization titer.

| | anti-N Ig ELISA | anti-N IgG ELISA | anti-S Ig ELISA | anti-S IgG ELISA | anti N/S Ig ELISA | anti N/S IgG ELISA |
|---|---|---|---|---|---|---|
| N LFRET | 88% (113/129) | 89% (115/129) | 91% (117/129) | 90% (116/129) | 87% (112/129) | 89% (115/129) |
| S LFRET | 93% (120/129) | 95% (122/129) | 96% (124/129) | 98% (127/129) | 92% (119/129) | 95% (122/129) |
| N/S LFRET | 95% (122/129) | 96% (124/129) | 98% (126/129) | 97% (125/129) | 94% (121/129) | 96% (124/129) |

**Table S2**. Detailed information on samples with discordance between PCR, LFRET and ELISA results. Sample = sample id. NAT = nucleic acid testing result for SARS-CoV-2. S = spike glycoprotein. N = nucleocapsid protein. LFRET = protein L–based time-resolved Förster resonance energy transfer immunoassay. ELISA = enzyme immunoassay. MNT = microneutralization titer. + = result above cutoff. - = result below cutoff. Hosp. = Hospitalized, non-ICU. ICU = Hospitalized in ICU. Home = non-hospitalized. Neg. = no disease. NA = not available.

| Sample | NAT | Severity of disease | Days from onset of symptoms | Days from NAT | anti-S LFRET | anti-N LFRET | anti-S ELISA IgA | anti-S ELISA IgG | anti-S ELISA IgM | anti-N ELISA IgA | anti-N ELISA IgG | anti-N ELISA IgM | MNT |
|---|---|---|---|---|---|---|---|---|---|---|---|---|---|
| 71 | + | Hosp. | 8 | 2 | - | - | - | - | - | + | - | - | <20 |
| 65 | + | ICU | 8 | 1 | - | - | - | - | - | - | - | - | <20 |
| 72 | + | ICU | 8 | 2 | - | - | - | - | - | - | - | - | <20 |
| 70 | + | ICU | 13 | 4 | - | - | - | - | - | + | - | - | 20 |
| 7 | + | Home | 30 | 24 | - | - | + | + | - | + | + | - | <20 |
| 24 | + | Home | 66 | 60 | - | - | - | + | - | + | + | - | <20 |
| 86 | + | Hosp. | NA | 2 | - | - | - | - | - | - | - | - | <20 |
| 82 | - | Neg. | 27 | 25 | - | - | - | - | - | - | + | - | NA |
| 92 | - | Neg. | 134 | NA | - | - | - | - | - | - | + | - | <20 |
| 103 | NA | Neg. | NA | NA | - | - | - | - | NA | NA | + | NA | <20 |
| 121 | NA | Neg. | NA | NA | - | - | + | - | NA | NA | - | NA | <20 |

**Table S3**. Sensitivity and specificity of LFRET/ELISA in detecting microneutralization positive (titer ≥ 20) samples. S = spike glycoprotein. N = nucleocapsid protein. LFRET = protein L–based time-resolved Förster resonance energy transfer immunoassay. ELISA = enzyme immunoassay. Only 4 negative samples were tested in anti-N IgA and IgM as well as anti-S IgM ELISA, and thus specificity of these assays needs to be assessed with caution.

| | anti-S LFRET | anti-N LFRET | anti-S IgA ELISA | anti-S IgG ELISA | anti-S IgM ELISA | anti-N IgA ELISA | anti-N IgG ELISA | anti-N IgM ELISA |
|---|---|---|---|---|---|---|---|---|
| Sensitivity | 98% | 92% | 98% | 98% | 79% | 88% | 98% | 20% |
| Specificity | 91% | 96% | 85% | 87% | 93% | 64% | 82% | 100% |
